# Social Isolation as a predictor for mortality: Implications for COVID-19 prognosis

**DOI:** 10.1101/2020.04.15.20066548

**Authors:** Sri Banerjee, Gary Burkholder, Beyan Sana, G. Mihalyi Szirony

## Abstract

The health benefits of social support have been widely documented. However, the social distancing practices from the COVID-19 pandemic is causing social disruption on a grand scale, potentially causing poor health outcomes. Through Google Trends analysis, we found a COVID-19-related surge in interest surrounding “loneliness.” We assessed if social isolation and loneliness increase the risk for all-cause and cardiovascular disease (CVD) mortality (ICD-10: I00–I99) and used the data to create a conceptual framework. Using the 10-year overall and cardiovascular mortality follow-up data (n = 12,019) from the National Health and Nutrition Examination Survey (1999–2008), we conducted survival analyses and found that individuals who experience social isolation or loneliness have a significantly higher likelihood of overall and CVD mortality than those without support. These effects generally remained strong with further adjustment for NHANES-detected health and demographic differences showing the need to address COVID-19 related loneliness through increasing social nearing.

## Introduction

The coronavirus disease (COVID-19) was the cause of a viral outbreak of respiratory illness in Wuhan, Hubei Province, China. As of 13 April 2020, this global pandemic had spread to 185 countries with 1.9 million confirmed cases, including 117,569 deaths, equating to a 6.2% global case fatality rate (Johns Hopkins University 2020). The magnitude, rapid rate of spread, and the high case fatality rate makes COVID-19 a global crisis (Adnan, Khan, Kazmi, Bashir, & Siddique 2020). Preventive measures such as masks, hand washing and hygiene practices, avoidance of public contact, avoidance of congregating in large crowds, increased interpersonal distance, and quarantines have been implemented as ways to reduce transmission in different states. To date, no specific antiviral treatment has proven effective. Therefore, infected people those infected primarily rely on symptomatic treatment and supportive care (Rodriguez-Morales et al. 2020). At the time of this writing, the COVID-19 pandemic is still spreading internationally and has had an enormous impact on human health, social life, and the economy. It also puts considerable strain on the healthcare system of nations worldwide.

Due to the COVID-19, pandemic social distancing has become the new norm of society. Social distancing has shown to be an effective way to slow down the spread of COVID-19, flattening the curve. Social distancing entails avoiding mass gatherings and maintaining a six-foot distance from other individuals has shown to be an effective way to slow down the spread of COVID-19 (IHME 2020). Using tenets from the field of proxemics, personal space which is reserved for close friends and family would be negatively affected due to the need for 1.5 to four feet. However, the objective of these social distancing measures has been to slow down the transmission of infection – or “flatten the curve” of the epidemic. Researchers have predicted that with social distancing measure in place, 1.7 million lives and $8 trillion will be saved by October 1 (Greenstone & Nigam 2020). The cancellations of large gathering, such as sporting events, cruises, musical events, festivals, and more, help stop or slow the spread of disease allowing healthcare facilities to care for patients over a period of time (New York Times 2020). The idea is to relieve the burden on healthcare systems in major cities and buy time until effective treatments and possibly a vaccine can be developed. However, as restrictions and regulations of social behavior are used as precautionary public health measures to prevent the spread of the pandemic, there is a risk of further marginalization and social isolation of individuals.

Loneliness has been considered an epidemic as well, as declared by previous world leaders (Klinenberg 2018). Current proportions are historically unprecedented at 60% or more in some modern European and North American cities (Snell 2017). Individuals who experience social isolation from social distancing risk experiencing loneliness. Social exclusion is particularly salient within older populations (Barbosa, Sanders, & Kokanovic 2019) that appear to be at the most risk for COVID-19 infection. Social pain is “the experience of social loss, situations in which valued social relationships and intimacy are threatened, harmed, or lost” (Baum, Lee, & Dougall 2011, p. 194). Thus, it refers to the unpleasant experience felt by actual or perceived damage to one’s sense of social connectedness or social value. Separation from loved ones, the loss of freedom, uncertainty over disease status, and boredom can be stressful resulting in harmful effects. Suicide and homicide have been reported, substantial anger generated, and lawsuits brought following the imposition of quarantine in previous outbreaks and the current pandemic (Sheperd 2020). Some researchers have coined the term ‘social recession’ to reflect the long-term consequences and impact of isolation and loss of human interaction (Murthy & Chen 2020).

While data is limited on the effects of quarantine and social isolation, there are studies that indicate psychological and physical health-related fallout due to quarantine. For instance, one study compared quarantined versus non-quarantined individuals during an equine influenza outbreak. Of 2,760 quarantined people, 34% reported high levels of psychological distress during the outbreak compared with 12% of non-quarantined individuals (Taylor et al. 2008). Brooks, Webster, Smith, Woodland, Wessley, Greenberg, and Rubin (2020), in a review of the literature reported frustration, boredom, post-traumatic stress, and anger, (p. 912) and acknowledged that some researchers postulated long-term harmful effects from quarantine. Social isolation has been found to be associated with depression, anxiety, and suicidal ideation. Further research suggests that physical and social pain might share biological substrates and extends this evidence base to the cardiovascular system (Inagaki et al. 2018). More specifically, increased blood pressure may serve as a regulatory function to modulate the effect of social pain, potentially affecting cardiovascular disease. However, other researchers have suggested from large representative samples that loneliness does not longitudinally result in cardiometabolic effects (Das 2018).

Although the connection between poverty, social determinants of health, social isolation, and poor health are well known, there is a paucity of research regarding the connection between social isolation and long-term health. Due to limited data from COVID-19, we decided to use nationally representative data to assess the effect of quarantining and social distancing on mortality. Specifically, in this study, we first explored how lonely people were feeling in the United States due to social distancing practices of COVID-19. We then explored the long-term associations between social isolation, loneliness, and all-cause and cardiovascular-related mortality. Furthermore, we used the findings from the research to conduct a conceptual framework for the societal effects of social distancing.

## Methods

We first used Google Trends to access Internet search patterns by analyzing a portion of all web queries on the Google Search website and other affiliated Google sites in the United States (Carneiro & Mylonakis 2009). We downloaded the output of their searches to conduct further analyses. We used the portal to determine the proportion of searches for a term “loneliness” over the time series of from 3/10/2019 to 4/5/2020 among all searches performed on Google Search, and found a relative search volume (RSV). The RSV is the query share of a particular term for a given location and time period, normalized by the highest query share of that. The reason we excluded any searches after April 5, due to the release of a popular song with term “loneliness” on April 7, which was released on this day.

Next, we used data from the National Health and Nutrition Examination Survey (NHANES), four cycles between 1999 to 2008. The analysis sample is representative of noninstitutionalized US adults, 20 years and older, to restrict the analysis to adults. This is representative of the adult population that is at risk from COVID-19 and related complications.

### Outcome Ascertainment

Vital status was determined using the Continuous NHANES Public-Use Linked Mortality File, which provides vital status follow-up data in person-months from the date of NHANES survey participation through the date of death or December 31, 2015. Mortality was ascertained by the NCHS through a probabilistic match between NHANES participants and National Death Index death certificate records. Participants who were not matched with death records were considered to be alive through the follow-up period. Cause of death was assigned by the NCHS based on the International Classification of Diseases, 10th Revision.

Cardiovascular mortality was specifically studied due to the fact that COVID-19 patients with cardiovascular disease are at a higher risk of mortality from these diseases (Clerkin et al. 2020). This strong connection may be due to the virus using the ACE-2 receptor, involved in blood pressure regulation, for host cell entry. For this study, cardiovascular mortality was defined as death due to diseases of the heart, essential hypertension and hypertensive kidney disease, cerebrovascular disease, atherosclerosis, and other diseases/disorders of the circulatory system (codes I00-I99).

### Social Isolation and Loneliness

Social isolation was measured by a single question, “Can you count on anyone to provide you with emotional support?” We focused on the emotional aspect of social support as this is especially affected during the COVID-19 pandemic and because prior empirical research has demonstrated that this is a key component of social support and is closely associated with depression.

Also, loneliness was considered a proxy for perceived inadequacy of emotional social support as has been well-defined in the literature (Perlman & Peplau 1981; Rico-Uribe et al., 2018). During COVID-19, many hospitals have completely restricted visitors for the inpatient population in hospitals due to concerns of viral spread. The emotional support is consequently restricted and potentially influencing health outcomes for even individuals who are not directly infected.

Perceived inadequacy was measured by respondents answering yes or no to a single item that inquired, “In the last 12 months, could you have used more emotional support than you received?” However, skip patterns designed into the NHANES interview protocol excluded this item when respondents had no one providing emotional support. We imputed a “yes” response for the respondents who skipped this item, and we conducted sensitivity analyses of the final model that revealed only a slight diminishment of effect. We analyzed this item separately from the social isolation indicator because of collinearity among the two indicators.

### Medical Covariates

The medical covariates are the conditions that were fitted into the same model as social isolation and loneliness. The following covariates were mainly included in the model as potential confounding variables.

#### CVD

Presence of CVD was determined by identifying those individuals that had a self-reported diagnosis of coronary heart disease, angina, stroke, congestive heart failure (CHF), or heart attack.

#### Obesity

Obesity was studied as well due to fact that obesity is a leading predictor of poor outcomes from COVID-19 (Dietz & Santos-Burgoa 2020). Obesity data were subdivided into four categories according to body mass index (BMI) derived from measured height and weight. The categories were as follows: participants with BMI < 25 were considered normal weight; participants with a BMI = 25-29 were overweight; participants with a BMI = 30-39.9 were considered as obese; and participants with a BMI > 40 were considered severely obese. For the multivariate models, obesity was dichotomized and considered present for BMI ≥ 30 and considered absent for the rest.

### Demographic Covariates

Education level, ethnicity, gender, and poverty level were tested for confounding effects. Gender was dichotomized as “male” and “female,” with female as the reference category. Education level was subdivided into a trichotomous indicator as “Completing some High School,” “High School graduate,” and “Some College or above.” Ethnicity was categorized as “Non-Hispanic White,” “Non-Hispanic Black,” “Hispanic,” and “Other.”

### Ethics Compliance

Before data collection for NHANES, the NCHS received approval from the NCHS Research Ethics Review Board (changed from the Institute Review Board (IRB)), continuance of the protocol #2011-17. The NCHS complies strictly with the different laws and regulations written with the intent of protecting the specific participant’s confidentiality and safety.

### Statistical Analysis

We compared means using the nonparametric Wilcoxon Mann-Whitney U test of the RSV from before and after 2/23/2020 in order to determine if COVID-19 in the United States had an effect on national search frequency.

Next, we weighted demographic and environmental variables to approximate distributions in the USA by using the provided sample weights to account for oversampling of young children, older individuals, Non-Hispanic Black individuals, and individuals of Mexican American ethnic origin in the NHANES survey. We adjusted for variables recognized widely as potential confounders for cardiovascular disease mortality. We adjusted all primary models for demographic and medical variables.

Statistical analyses were conducted using the SAS System for Windows (release 9.1; SAS Institute Inc., Cary, NC) and SUDAAN (release 9.0; Research Triangle Institute, Research Triangle Park, NC). Weighted Cox proportional hazard regression models were used to examine the relationship between social isolation/loneliness and total and cardiovascular mortality. All analyses included sample weights that accounted for the unequal probabilities of selection and nonresponse. All variance calculations incorporated the sample weights and accounted for the complex sample design using Taylor series linearization. All significance tests were two-sided using p < 0.05 as the level of statistical significance.

## Results

Our Google Trends search data shows that the relative search volume for “loneliness” has been the highest since when Google Trends started measuring trends in 2004. As seen in Figure 1, when considering the term “loneliness” from 3/10/2019 to 4/5/2020, the relative search volume has been increasing since February and highest during the time period. In comparison to March and April of 2019, loneliness was experienced much more acutely in the United States. Temporally, the RSV mean (25.5 vs. 54, p < 0.001) was statistically significantly different statistically for before and after 2/23/2020. This date is in the middle of the time interval between when COVID-19 was first recognized in the United States in Washington State and the date that it was first found in New York State. This has coincided with the COVID-19 pandemic and implementation of social distancing policies that had been put in place varying widely throughout the US.

**Figure 1.**
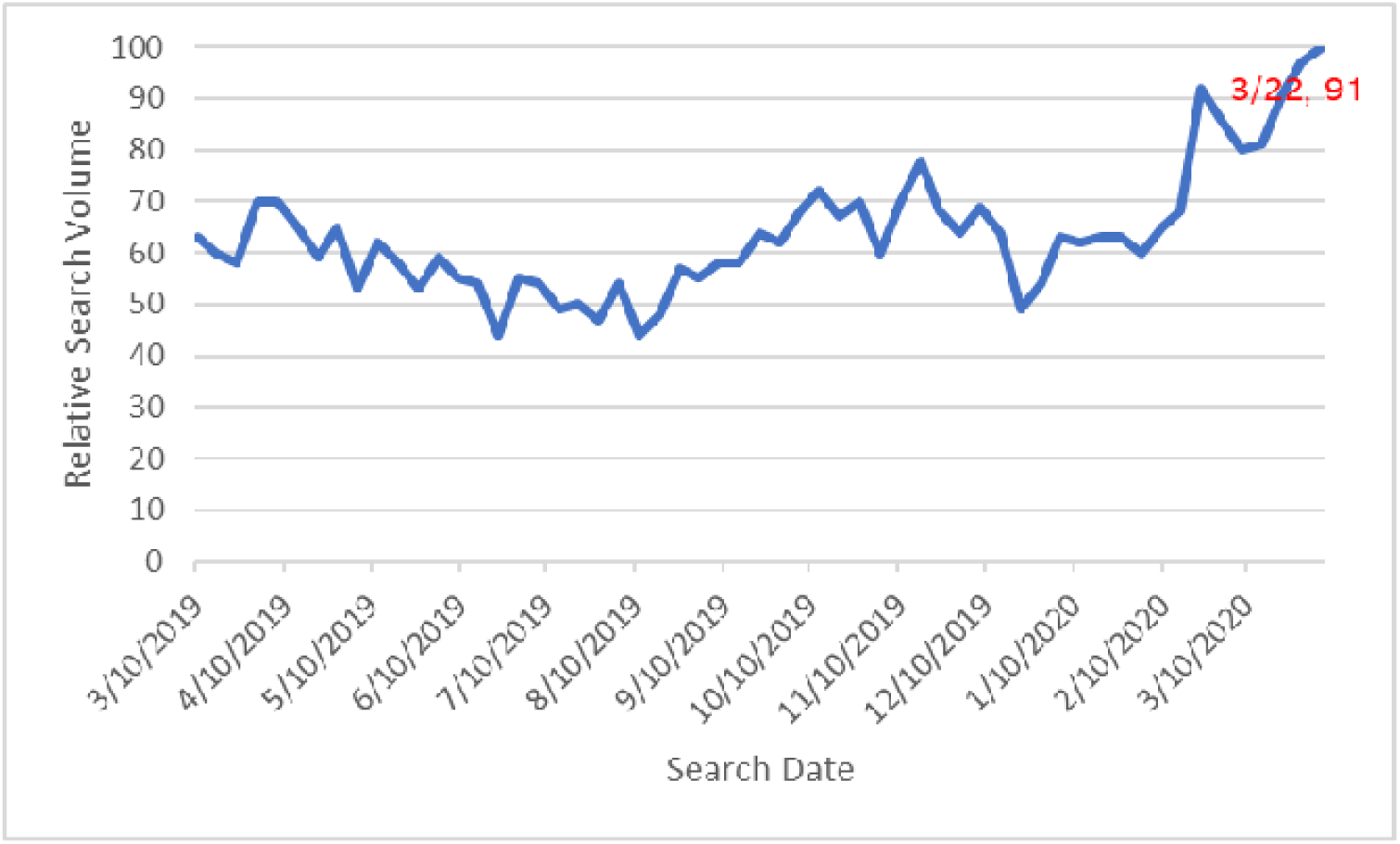
Relative Search Volume for the search term “Loneliness” in the United States

### NHANES Analysis

Our analysis of NHANES data included 12,954 subjects aged 20 years or greater. Table 1 provides the data for the distribution of the demographic characteristics and medical risk factors of the participants by the status of social support using bivariate analysis. The weighted prevalence of loneliness in the US population within the 20 years or older age group was 23.2% (95% CI=22.1-24.3), which is representative of 22,213,619 individuals (males: 42.6% vs. females: 57.4%, p<0.001) in the United States population. Also, the weighted prevalence of social isolation in the US population within the 20 years or older age group was 5.6% (95% CI=5.0-6.2), which is representative of 5,309,054 individuals (males: 50.9% vs. females: 49.1%, p=.03) in the United States population. The average age of the participants within the sample was 46.4 ± 0.24 years. There was a statistically significant (P < 0.05) association between age, gender, race/ethnicity, education level, CVD status, obesity status, diabetes and social isolation. As shown in Table 1, those individuals that are socially isolated are slightly older (60.3 vs. 59.6) in age than those that have social support. Among those individuals who were socially isolated, participants were more likely to be a minority (38.5%), with less than a high school education (35.9%), and people diagnosed with cardiovascular disease (19.7%) compared to their counterparts who did not report social isolation and were considered having social support (more likely to be Non-Hispanic White, with higher levels of education, and less likely to have CVD).

**Table 1.** Mortality frequency and percentage of Study Participants^a^ stratified by Social Support Status

| Variable | Total population<br>(n=12,954) | Social Support<br>(n=11,960) | Social Isolation<br>(n=994) |
| --- | --- | --- | --- |
| **All Deaths** | 3972 (22.7%) | 3648 (22.4%) | 324 (27.9%) |
| **Cardiovascular Disease Deaths** | 743 (17.8%) | 672 (17.4%) | 71 (23.0%) |
| **Demographic Risk Factors** |  |  |  |
| **Female** | 6577 (53.9%) | 6116 (54.1%) | 461 (49.1%) |
| **Mean age (SE)** | 46.4 (0.24) | 59.6 (0.27) | 60.3 (0.56) |
| **Ethnicity\*\*** |  |  |  |
| **Non-Hisp. White** | 6994 (77.0%) | 6599 (77.9%) | 395 (61.5%) |
| **Non-Hisp. Black** | 2564 (10.0%) | 2403 (9.9%) | 161 (11.0%) |
| **Hispanic** | 2975 (8.4%) | 2569 (7.7%) | 406 (20.0%) |
| **Other** | 421 (4.5%) | 389 (4.4%) | 32 (7.5%) |
| **Education Level\*\*** |  |  |  |
| **Some High School** | 4178 (21.0%) | 3969 (20.1%) | 482 (35.9%) |
| **High School Grad** | 3090 (26.8%) | 2884 (26.8%) | 206 (26.1%) |
| **Some College and beyond** | 5366 (52.2%) | 5097 (53.0%) | 269 (38.0%) |
| **Medical Risk Factors** |  |  |  |
| **Cardiovascular Disease** | 2547 (16.0%) | 2352 (15.8%) | 195 (19.7%) |
| **Obesity Status\*\*** |  |  |  |
| **Normal Weight**<br>**BMI < 25** | 3353 (28.3%) | 3107 (28.5%) | 246 (25.8%) |
| **Overweight**<br>**BMI = 25 – 29.9** | 4560 (36.1%) | 4191 (36.0%) | 369 (39.0%) |
| **Obese**<br>**BMI = 30—39.9** | 3764 (30.2%) | 3486 (30.2%) | 278 (29.7%) |
| **Morbidly Obese**<br>**BMI ≥ 40** | 640 (5.4%) | 596 (5.4%) | 44 (5.4%) |

Note. \* $p < .05$ \*\* $p < .001$
<sup>a</sup> Data are number, %, or mean. Percentages and means are weighted to match the age, sex, and ethnic origin distribution of the US population.

### Mortality

Out of 994 participants (54% females vs. 46% males) with social isolation, 324 deaths were reported (including 71 CVD deaths) during an average of 10-year follow-up. The unadjusted hazard ratio for overall mortality among those experiencing social isolation was 1.35 (95% CI = 1.18-1.54). The age- and gender-adjusted hazard ratio for CVD-mortality among those experiencing social isolation was 1.44 (95% CI = 1.08-1.92). The hazard ratio for age- and gender-adjusted mortality among those experiencing loneliness compared with those not lonely is 1.28 (95% CI = 1.17-1.39). The age- and gender-adjusted hazard ratio for CVD-mortality among those experiencing loneliness was 1.36 (95% CI = 1.15-1.61).

### Adjusted Loneliness and Social isolation Mortality

When considering social isolation, the adjusted HR for all-cause mortality [1.20 (CI 1.04-1.38, p = 0.01)] was significant, after additional adjustment for demographic (age, gender, and ethnicity) and health risk factors (cardiovascular disease and obesity), as seen in table 2. Finally, the adjusted HR for cardiovascular mortality [1.44 (CI 1.04-2.01, p = 0.03)] remained significant, after adjusting for age, gender, ethnicity, education, pre-existing cardiovascular disease, and obesity.

**Table 2.** Multivariable Cox Hazard Model for social isolation and all-cause mortality (n=11,919) or CVD-mortality (n=3,450) after controlling for demographic and medical risk factors.

| Variable | All-Cause Mortality |  |  | CVD-Mortality |  |  |
| --- | --- | --- | --- | --- | --- | --- |
|  | HR | 95% CI | P-value | HR | 95% CI | P-value |
| Social Isolation | 1.20 | 1.04-1.38 | 0.01 | 1.44 | 1.04-2.01 | 0.03 |
| Cardiovascular Disease<sup>a</sup> | 1.91 | 1.74-2.10 | <0.001 | 1.90 | 1.53-2.36 | <0.001 |
| Obesity<br>(Reference: BMI < 30) | 1.23 | 1.12-1.34 | <0.001 | 1.16 | 0.89-1.50 | 0.26 |
| Education |  |  |  |  |  |  |
| Some High School | 1.38 | 1.26-1.50 | <0.001 | 1.04 | 0.80-1.35 | 0.78 |
| High School Grad | 1.26 | 1.13-1.40 | 0.002 | 1.12 | 0.89-1.41 | 0.002 |
| Some College | Reference | Reference | Reference | Reference | Reference | Reference |
| Age | 1.09 | 1.09-1.10 | <0.001 | 1.00 | 0.99-1.02 | 0.71 |
| Gender<br>(Reference: Female) | 1.42 | 1.28-1.57 | <0.001 | 1.49 | 1.24-1.79 | <0.001 |
| Ethnicity |  |  |  |  |  |  |
| Non-Hispanic White | Reference | Reference | Reference | Reference | Reference | Reference |
| Non-Hispanic Black | 1.27 | 1.14-1.41 | <0.001 | 1.35 | 1.05-1.74 | 0.02 |
| Hispanic | 0.97 | 0.84-1.12 | 0.68 | 1.18 | 0.84-1.66 | 0.33 |
| Other | 1.02 | 0.75-1.39 | 0.90 | 1.05 | 0.56-1.98 | 0.88 |

<sup>a</sup>Cardiovascular disease was defined by self-reported positive response to congestive heart failure, stroke, angina, coronary heart disease, or heart attack.

When considering loneliness, the adjusted HR for all-cause mortality [1.24 (CI 1.12-1.38, p = 0.01)] was significant, after additional adjustment for demographic (age, gender, and ethnicity) and health risk factors (cardiovascular disease and obesity), as seen in Table 3. Finally, the adjusted HR for cardiovascular mortality [1.31 (CI 1.09-1.56, p < 0.05)] remained significant, after adjustment for age, gender, ethnicity, education, pre-existing cardiovascular disease, and obesity.

**Table 3.** Multivariable Cox Hazard Model for loneliness and all-cause mortality (n=12,019) or CVD-mortality (n=3,485) after controlling for demographic and medical risk factors.

| Variable | All-Cause Mortality |  |  | CVD-Mortality |  |  |
| --- | --- | --- | --- | --- | --- | --- |
|  | HR | 95% CI | P-value | HR | 95% CI | P-value |
| Loneliness | 1.24 | 1.12-1.38 | <0.001 | 1.31 | 1.09-1.56 | <0.05 |
| Cardiovascular Disease<sup>a</sup> | 1.89 | 1.72-2.07 | <0.001 | 1.91 | 1.54-2.37 | <0.001 |
| Obesity<br>(Reference: BMI < 30) | 1.23 | 1.13-1.35 | <0.001 | 1.15 | 0.88-1.49 | 0.31 |
| Education |  |  |  |  |  |  |
| Some High School | 1.37 | 1.25-1.49 | <0.001 | 1.06 | 0.82-1.36 | 0.67 |
| High School Grad | 1.26 | 1.12-1.41 | <0.001 | 1.12 | 0.90-1.39 | 0.32 |
| Some College | Reference | Reference | Reference | Reference | Reference | Reference |
| Age | 1.09 | 1.09-1.10 | <0.001 | 1.00 | 0.99-1.02 | 0.63 |
| Gender<br>(Reference: Female) | 1.44 | 1.30-1.58 | <0.001 | 1.54 | 1.27-1.85 | <0.001 |
| Ethnicity |  |  |  |  |  |  |
| Non-Hispanic White | Reference | Reference | Reference | Reference | Reference | Reference |
| Non-Hispanic Black | 1.26 | 1.12-1.41 | <0.001 | 1.34 | 1.05-1.72 | 0.02 |
| Hispanic | 0.95 | 0.82-1.09 | 0.25 | 1.15 | 0.82-1.60 | 0.25 |
| Other | 1.00 | 0.73-1.37 | 0.99 | 1.06 | 0.56-2.03 | 0.85 |

<sup>a</sup>Cardiovascular disease was defined by self-reported positive response to congestive heart failure, stroke, angina, coronary heart disease, or heart attack.

Using the evidence from the research on loneliness and social isolation, as seen in Figure 2, we created a Social Distancing Conceptual Model (SDCM) in order to demonstrate the potential trajectories of how a person reacts to the policies implemented to curb the spread of COVID-19. As is demonstrated, social isolation is not automatically the end result of social distancing. The left part of the model is policies related to COVID-19, and the right is the long-term outcomes upon members of society. The top half of the model demonstrates that individuals who are flexible and are well adjusted can adapt to particular technologies to aid in social nearing. This ability to adapt has as much to do with adaptability as has to do with individuals who are not socially oppressed. For instance, ethnic minorities face more discrimination and stress than those individuals of the majority population (e.g, Williams 2018; Wong-Padoongpatt, Xane, Okazaki, & Saw 2020). As individuals feel supported due to the effective usage of video chatting technology, they can also feel a sense of social responsibility—moving along from left to right on the top segment of the model. By understanding the importance of social responsibility, this can lower mortality and create a sense of social cohesion. Also, this can leadto bridging social capital in that bonds can be created between ethnic groups and other social cleavages. This leads to the norm of reciprocity loop which is the expectation that people will respond favorably by returning benefit to benefit (Kjørstad 2017). This loop allows for the building of social support and continuous positive outcomes for the individual and society. However, if the individual feels stigmatized and feels disempowered, the individual can then switch from the top half of the model to the bottom half.

**Figure 2:**
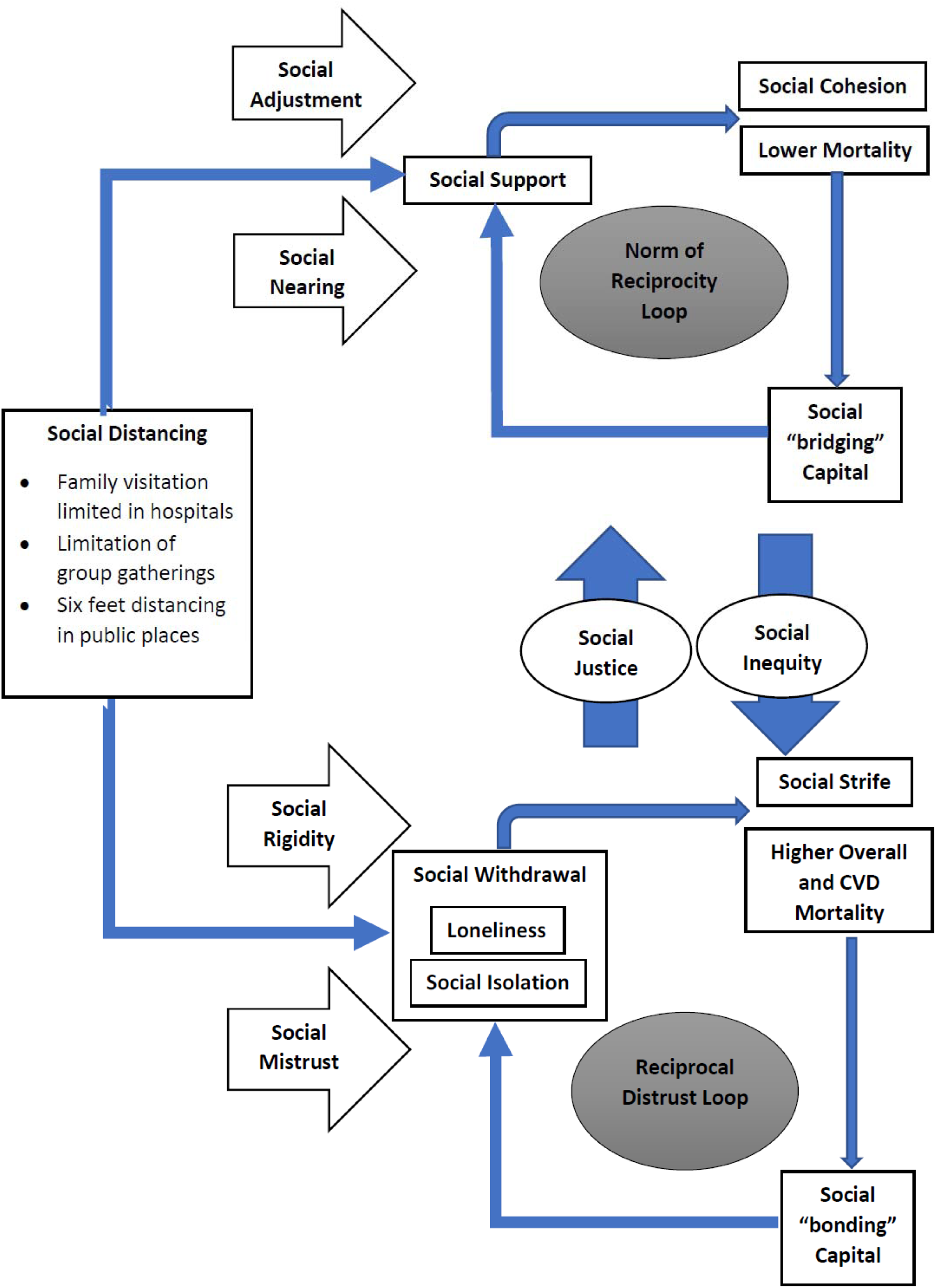
Social Distancing Conceptual Model (SDCM) used to inform potential trajectories of Social Distancing Policies

As demonstrated in the SDCM, not all individuals socially distancing, will feel supported. For instance, people who are socially marginalized and experience racism may have feelings of social mistrust and social rigidity. Many times, the elders or ethnic minorities feel socially withdrawn. These social distancing rules may result in certain individuals experiencing loneliness and social isolation. This may lead to an increase in overall and CVD mortality as is evidenced by our research. This may also lead to social strife due to a feeling of resentment. A maladaptive result may be bonding social capital which is exclusive bonding only within a social group. The person may look to blame minority groups for the spread of disease or feel isolated or stigmatized if they have the disease. This viewpoint would lead to further social withdrawal and the perpetuation of violence and strife. This leads to an iterative loop which can lead to social disintegration as has been evidenced by previous researchers (Gorman 2005).

## Discussion

The findings from this study help highlight the loneliness experienced by the populace, due to the mode of transmission and policies resulting from COVID-19. The upsurge of popularity of “loneliness” in Google Trends directly coincided with the timeline of events surrounding COVID-19. For instance, after the first case in January 20, 2020 in Washington, many governments in different areas started considering social distancing policies (Rothan & Byareddy 2020). Fear of transmission and infection further led to isolating behaviors like avoidance of large crowds. The main areas that they considered were the closure of educational facilities, a stay at home order, closure of non-essential businesses, and limitation of travel (IHME 2020).

All of these policies were potential contributors of loneliness. As expected, one of the major drivers of how loneliness was experienced in the United States is how people experienced this feeling in New York City. On March 18, 2020, New York’s governor closed down schools (IHME 2020). Subsequently, by March 22, 2020, the state governor implemented a stay at home order for the state. This can be clearly linked to a relative search volume of 91 for the term “loneliness.”

The findings from this study help demonstrate a clear link between loneliness and social isolation, as experienced due to COVID-19 related policy, and mortality even aside from the presence of other important determinants of mortality. Among adults, social isolation is associated with all-cause mortality and cardiovascular mortality. According to our findings, social isolation is associated with 35 percent higher overall mortality rate than those individuals who have social support. Previously, Holt-Lunstad et al. (2015) conducted a meta-analysis of 70 studies involving more than 3.4 million participants followed for an average of seven years. The likelihood of dying during the study period increased by 26 percent for those who reported loneliness (feeling alone), 29 percent for those who were socially isolated (having few social contacts) and 32 percent for those living alone.

According to our study, national loneliness rates were found to be 23 percent with females statistically significantly experiencing loneliness at higher rates than males. In the context of COVID-19, loneliness would be experienced at higher rates due to stay-at-home orders and restricted visitor policies at hospitals (New York Presbyterian Hospital 2020). Patients, even those not directly infected by COVID-19, are dying alone and loved ones cannot grieve in the traditional way since funeral gatherings are banned (Willis & Williams 2020). Gender disparities were not as striking in social isolation rates. Previous studies found loneliness rates of 17 percent to 57 percent (Lee et al. 2019). Unlike our study, men and women were equally affected by loneliness in previous studies.

Another novel finding from our study is that loneliness is considered an important risk factor of mortality. According to our findings, loneliness is associated with 28 percent higher overall mortality rate than those individuals who did not experience loneliness. This association remained strong even after controlling for medical and demographic health variables. This finding is also consistent with our findings of social support.

Other studies have provided evidence of the relationship between human touch, social cohesion, and physiological well-being. Modern technology cannot substitute human touch, such as holding hands, hugging, or massage, which studies suggest can affect health, including possibly lowering blood pressure and reducing the severity of symptoms from the common cold. Social contact has been connected to the release of oxytocin (Holt-Lunstad et al. 2008). Loneliness has been associated with all-cause mortality in a previous meta-analysis study (Rico-Uribe et al., 2018). Specifically, it was found that individuals who have an anxious attachment style have an increased neurological sensitivity to social distress. This is expected due to the fact such individuals crave acceptance and look to external cues for any chance of rejection. However, in the absence of the possibility of social contact due to threat of viral contraction, other strategies must be employed in order to maintain social connectedness, even in those with an anxious attachment style.

Our SDCM model is the first conceptual model based on COVID-19 to date. This model can be used to mitigate the effect of social distancing by helping identify how loneliness leads to higher rates of mortality and how social disparities help guide how the person will be impacted from social distancing. Initial evidence certainly supports the fact that social distancing will negatively affect minorities more so than Caucasians (Rundle et al., 2020). The existing racial disparities and social determinants of health are amplified by the effect of COVID-19. Furthermore, negative adjustment to social distancing perpetuates violence. News media is replete with examples of individuals buying firearms and using these in domestic disputes after the COVID-19 crisis. For instance, a Pennsylvania man shot his girlfriend and killed himself over COVID-19 worries and loss of employment (Burke 2020). A similar murder suicide also took place in another state due to the worry that the couple had COVID-19 (Sheperd 2020). However, after death, they both were found to be free of the virus. If the social injustices are addressed through policies like economic stimulus plans to benefit disenfranchised members of society, then the deleterious effects of loneliness and social isolation may be mitigated.

Human interaction promotes the release of endorphins and oxytocin. Without interaction and support, there is a feeling of loneliness and isolation which can have negative consequences on the individual. Social isolation and lack of social support are likely acute and chronic stressors affecting biological and behavioral mediators, such as increasing allostatic overload or unhealthy behaviors (Zeilowsky et al. 2018). Such mediating pathways are postulated to have long-term negative effects on health, causing increases in disease susceptibility and risk of mortality across many leading causes of death among elders. The role of social disconnectedness is particularly salient among populations with greater susceptibility to morbidity and mortality, such as older adults. The lack of social support for this population incurs real societal costs, such as longer hospital or nursing home stays when older persons lack caregivers who can help them recover at home (Taylor et al. 2008). Additionally, prolonged isolation can lead to hyperarousal and hypervigilance, leading to further depression or violence.

### Implications for Practice

Social distancing, as recommended due to COVID-19, can lead to feelings of social isolation. Efforts should be taken to mitigate effects of social distancing. One method is to practice social nearing while maintaining social distancing. This can be accomplished by virtually utilizing video chat apps and optimally using texting capabilities, the use of which is being encouraged by some hospitals (Penn Medicine 2020). Family support, which is restricted due to COVID-19, is typically an integral part of treatment within hospitals (New York Presbyterian Hospital 2020). Borrowing from social psychology and computer-mediated communication research, there are certain strategies that can be used to increase social presence by family members and create an atmosphere of support for the patient (Dixson et al. 2015). The concept of social presence can be used to classify multimedia interfaces according to how well they convey intimacy and warmth between users. Specifically, during communication, using emoticons and figurative language helped to increase social presence and immediacy, which is defined by nonverbal and verbal cues that can be used to decrease psychological distancing and increase closeness (Schultze & Brooks 2019). When texting, in order to minimize social isolation and psychological distancing, family members should look to decreasing response latency in order to make a difference in health outcomes. Minimizing the lag time between initial message and response can help improve immediacy.

Also, social isolation and loneliness factors should be considered an independent social determinant and a risk factor for CVD in clinician risk scoring models. Including social isolation into risk scoring models can lead to better classification of those at high risk of CVD. Health practitioners should consider social isolation, along with other social determinants of health, when determining an individual’s overall health risk and especially their risk for cardiovascular disease. As Brooks et al. (2020) noted, that public health officials should exercise caution and quarantine and isolate only when absolutely necessary to minimize the effects of social isolation. Also, strong negative words like “prohibited” in hospital visitor policies should be discouraged, as this creates a norm of social isolation. Instead, researchers from a randomized controlled trial of social distancing communications found that instructions with information on transmissibility and transmission rate is less effective than communications which entail the potential transmission to identifiable family members. People are able to better connect and identify with social distancing scenarios illustrating transmission to family members (Lunn et al. 2020).

### Recommendations for Future Studies

More research is needed in determining best practices for addressing social isolation in an effective, efficient, and dignified manner, especially for those directly and indirectly affected by COVID-19 policy. Also, more research is need in determining effective interventions for individuals who are in social isolation.

## Data Availability

All data is freely available through Google and CDC.

https://trends.google.com/trends/?geo=US

https://wwwn.cdc.gov/nchs/nhanes/

## Authors’ contributions

SB, GB, BS, and GS conducted the literature review and contributed to the writing of the manuscript. SB conducted all statistical analyses and SB and GB contributed to the design of the study. GB, BS, and GS provided critical feedback on the study design and results interpretation.

## Availability of data and materials

The datasets used and/or analyzed during the current study are available from the Centers for Disease Control website https://wwwn.cdc.gov/nchs/nhanes/Default.aspx.

## Declaration of conflicting interests

The authors declared no potential conflicts of interest with respect to the research, authorship, and/or publication of this article.

## Funding

The authors received no financial support for the research, authorship, and/or publication of this article.

## Ethical Standards Disclosure

This study was conducted according to the guidelines laid down in the Declaration of Helsinki and all procedures involving research study participants were approved by the NCHS Research Ethics Review Board and Walden University Institutional Review Board. Written informed consent was obtained from all subjects.

## Notes

### Competing Interest Statement

The authors have declared no competing interest.

### Funding Statement

None

